# Home Monitoring for Fever: An Inexpensive Screening Method to Prevent Household Spread of COVID-19

**DOI:** 10.1101/2021.12.21.21268203

**Authors:** Justin Kim, Marcus A. Threadcraft, Wei Xue, Sijie Yue, Richard P. Wenzel, Frederick S. Southwick

**Author notes:** Corresponding Author, 6362 NW 41^st^ Ave, Gainesville, FL 32606.

## Abstract

The COVID-19 pandemic surge has exceeded testing capacities in many parts of the world. We investigated the effectiveness of home temperature monitoring for early identification of COVID-19 patients.

**Study Design:** We compared home temperature measurements from a convenience sample of 1180 individuals who reported being test positive for SARS-CoV-2 to an age, sex, and location matched control group of 1249 individuals who had not tested positive.

**Methods:** All individuals monitored their temperature at home using an electronic smartphone thermometer that relayed temperature measurements and symptoms to a centralized cloud based, de-identified data bank.

**Results:** Individuals varied in the number of times they monitored their temperature. When temperature was monitored for over 72 hours fever (> 37.6°C or 99.7°F or a change in temperature of > 1°C or 1.8°F) was detected in 73% of test positive individuals, a sensitivity comparable to rapid SARS-CoV-2 antigen tests. When compared our control group the specificity of fever for COVID-19 was 0.70. However, when fever was combined with complaint**s** of loss of taste and smell, difficulty breathing, fatigue, chills, diarrhea, or stuffy nose the odds ratio of having COVID-19 was sufficiently high as to obviate the need to employ RTPCR or antigen testing to screen for and isolate coronavirus infected cases.

**Conclusions:** Our findings suggest that home temperature monitoring could serve as an inexpensive convenient screen for the onset of COVID-19, encourage earlier isolation of potentially infected individuals, and more effectively reduce the spread of infection in closed spaces.

## Introduction

In many regions of the world the COVID-19 pandemic remains poorly controlled and vaccines supplies are limited; therefore, nonpharmacologic approaches to infection control will continue to play a central role in reducing infection spread. Testing capacity is limited in many parts of the world and the results from RTPCR tests may take days to return.^1,2^ These conditions delay rapid identification, isolation, contact tracing, and early treatment of those who are infected by SARS-CoV-2.^3^ Eighty percent of new cases are contracted in the household. Secondary attack rates following identification of an index case within the household vary from 6.7 to 31.3% ^4^ and with the spread of the more contagious delta variant secondary household attack rates are likely become even higher. Infections within the household can spread quickly, 75% of household cases contracting the infection within 5 days of the index case in one study, emphasizing the importance of early screening and isolation.^5^ Is there a better way to quickly identify the onset of disease and encourage earlier isolation of potentially SARS-CoV-2 infected individuals?

To explore the potential utility of home monitoring for fever as a preliminary screening tool, we contacted the smartphone thermometer company Kinsa Inc. that provided us with a national convenience sample of temperature measurement from 1180 individuals who reported testing positive for SARS-CoV-2 and had monitored their temperatures at home.

Our analysis reveals that home monitoring of core temperature for over 3 days detects fever in COVID positive patients, with sensitivities comparable to RTPCR and rapid antigen testing. Logistic regression analysis combining the presence of fever with individual symptoms generated odds ratios that can guide the selection of family members who should be isolated and tested.

## Methods

Kinsa Smart oral and ear thermometers (https://www.kinsahealth.co/) record and store temperature measurements using the Kinsa smartphone application.^6^ With the acknowledged agreement by the users, temperatures are downloaded to a national data base that also includes the location of the measurements using GPS coordinates, age, gender, dates and times of measurements, measurement site (oral, axillary, ear, or rectal), and a symptom checklist.

### Study design

The data provided by Kinsa Inc represented a national convenience sample, that was deidentified and provided in raw form in an Excel format as previously described.^6^ Our study was approved by the University of Florida IRB as exempt (UF IRB# 202003028).

### Participants

The sample included 1180 individuals ages 2 years and older who reported testing positive for SARS-CoV-2 and 1249 control individuals matched for location, gender and age who had not reported a positive test. A total of 38,901 temperature measurements were analyzed for the COVID-19 group and 37,420 measurements for the control group over a 10-month period from February 21 to December 20, 2020.

### Test methods

Fever was defined as a core temperature of > 37.6°C (72% of fevers in our series) or a change in temperature of >1°C (28%) over the period of monitoring. There is a broad range of published estimates for a normal oral temperature, however, based on the conclusions of a systematic review of these studies^7^ we chose a mean oral temperature of 36.6°C as best reflected the mean normal temperature for all age groups. The definition of what constitutes fever also varies, and among studies examining fever in COVID-19 patients fever cutoffs ranged from >37.2°C ^8^ to > 38°C^9^. We chose a value of 37.6° midway between these two extremes and 1 °C ° above the chosen normal temperature. Recognizing that the thermal set point decreases as individuals age, we also used a rise in temperature of 1°C ° during the observation period as a second definition for fever. For the SARS-CoV-2 positive group, we collected temperature measurements beginning 14 days prior to the date of the positive test, and 14 days afterwards. Fourteen days was chosen because this is incubation period for COVID-19 determined by the CDC. For the matching control group, we analyzed temperatures for 28 days during the same period counting back from the most recent temperature measurement.

In addition to recording their temperatures participants were also prompted by the app to check off any symptoms they were experiencing, and we assessed the sensitivity and specificity of each individual symptom as well as applied step wise logistic regression analysis to determine if fever and specific symptoms were independent predictors of having Covid-19.

### Analysis

A two-sided Fisher’s exact test was applied using contingency tables to assess the statistical significance of differences in the frequency of fever and individual symptoms between SARS-CoV-2 positive and neg individuals, as well as to determine the sensitivities and specificities of each variable. These analyses were performed using Prism 9.0 by GraphPad LLC.

To test the accuracy of using fever as main measurement of predicting SARS-CoV-2, several logistic regression models were implemented adjusting for different symptoms. The null model of defining the association of fever and SARS-CoV-2 was fitted. Then the logistic regression models of fever and loss of taste or smell, fever and cough, fever and trouble breathing, fever and stuffy nose, fever and headache, fever and chills, fever and diarrhea, fever and fatigue, and fever and body ache were fitted separately. Sensitivity of each logistic regression models was calculated to compare the results. P-values less than 0.05 were deemed statistically significant. R version 4.0.2 was utilized for all the statistical modelling work.

## Results

### Participants

Forty-six states excluding Hawaii, Maine, New Mexico, and Wyoming were represented in the sample and reflected the relative percentages of the U.S. population for each state with the exceptions of underrepresentation of Florida (2.7 vs 6.5%) and overrepresentation of Kansas (3.2 vs 0.9%), Connecticut (3.4 vs 1.1%), and Minnesota (3.8 vs 1.7%). Females were overrepresented, being 63% of users. Age distribution approximated that of the U.S. with the exception of over representation of ages 19-60 years (72 vs 53%) and underrepresentation of ages >60 years (7.2 vs 22%). Most temperature measurements were oral (97%). The number of daily new case of SARS-CoV-2 infection in the U.S. varied during the period varied from <1/100,000 in February. 2020 to 54/100,000 in November and 65/100,000 in December 2020.

### Test Results

Fever was present in 63.1% of all SARS-CoV-2 test positive individuals. (Table 1). A single temperature check was insensitive, detecting fever in only 30.3% of cases. To determine the duration of temperature monitoring that maximized detection of fever we plotted percent with fever versus the duration of temperature monitoring (Figure 1). The percent with fever plateaued between 72 and 96 hours. Subsequent analysis of all SARS-CoV-2 positive patients who monitored their temperature over 72 hours revealed that 73.1% had fever. Using the definitions of fever described in the methods we found the percentage with fever did not vary significantly by age group.

**Figure 1.**
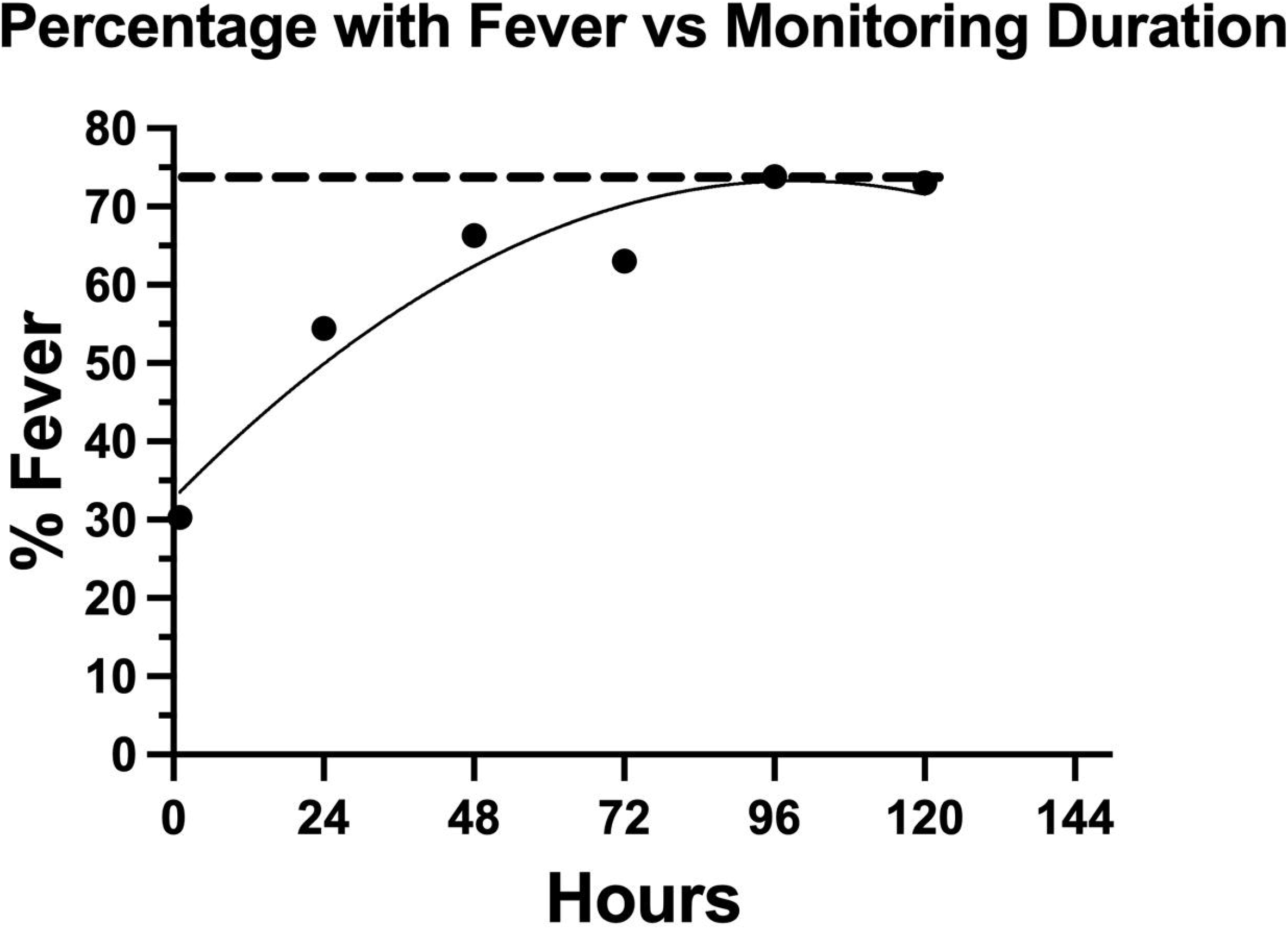
Symptoms Associated with COVID-19. 1081 patients who tested positive for SARS-CoV-2 monitored their oral temperature at home for differing durations. This graph shows the relationship between duration of temperature monitoring and detection of fever. The percentage with fever plateaued between 72 and 96 hours at 74%.

**Table 1.** Presence of fever in individuals testing positive for SARS-CoV-2.

| Age group | % Fever Total<br>(#fever /total) | % Fever Single<br>(#fever/total) | % Fever <72 h<br>(#fever/total) | % Fever >72 h<br>(#fever/total) |
| --- | --- | --- | --- | --- |
| 2-6 y | 69.2% (36/52) | 62.5% (10/16) | 75.0% (12/16) | 70.0% (14/20) |
| 7-12 y | 65.4% (53/81) | 40.0% (8/20) | 67.9% (19/28) | 78.8% (26/33) |
| 13-18 y | 65.1% (56/86) | 61.9% (13/21) | 58.6% (17/29) | 72.2% (26/36) |
| 19-60 y | 62.4% (547/876) | 19.6% (22/112) | 59.8% (140/234) | 72.6% (385/530) |
| > 60 y | 62.4% (53/85) | 18.8% (3/16) | 54.6% (6/11) | 75.9% (44/58) |
| All ages | 63.1% (745/1180) | 30.3% (56/185) | 61.0% (194/318) | 73.1% (495/677) |

To assess the specificity of fever we compared the incidence of fever to an age, gender, geographically matched group who had monitored their temperature during the same period and did not report testing positive for SARSCoV-2. (Table 2) Fever was present in 35.6% of controls. When temperature was monitored for > 72 hours fever was detected in 42.2% of cases.

**Table 2.** Presence of fever in individuals not testing positive for SARS-CoV-2.

| Age group | % Fever Total<br>(#fever /total) | % Fever Single<br>(#fever/total) | % Fever <72 h<br>(#fever/total) | % Fever >72 h<br>(#fever/total) |
| --- | --- | --- | --- | --- |
| 2-6 y | 53.7% (22/41) | 30.8% (4/13) | 64.3% (9/14) | 64.3% (9/14) |
| 7-12 y | 46.2% (36/78) | 9.5% (2/21) | 64.7% (11/17) | 57.5% (23/40) |
| 13-18 y | 35.4% (28/79) | 13.0% (3/23) | 50.0% (8/16) | 42.5% (17/40) |
| 19-60 y | 33.0% (315/955) | 9.7% (22/226) | 43.9% (76/173) | 39.0% (217/556) |
| > 60 y | 45.8% (44/96) | 16.7% (2/12) | 25.0% (2/8) | 52.6% (40/76) |
| All ages | 35.6% (445/1249) | 11.2% (33/295) | 46.5% (106/228) | 42.2% (306/726) |

Contingency analysis revealed that the specificity of fever for COVID-19 in all patients who checked their temperature was 0.649 (range 0.622 to 0.675) (p< 0.0001). The positive predictive value was 0.631 (range 0.6034 to 0.6584), the negative predictive value 0.644 (0.617 to 0.670). For those who monitored their temperature for > 72 hours specificity increased to 0.698 (0.660 to 0.733) (p < 0.0001), the positive predictive value was 0.727 (0.692 to 0.760), the negative predictive value 0.578 (0.542 to 0.614),

In addition to fever 84.6% of patients who tested positive for SARS-CoV-2 and 72.5% of those who had not tested positive reported symptoms at the time of temperature monitoring. Fever and symptoms occurred within 12 hours of each other in 453 of 661 febrile symptomatic test positive individuals (68.5%). Fever developed greater than 12 hours before symptoms in 127 cases (19.2%) and symptoms preceded fever by more than 12 hours in only 81 cases (12.2%). Overall fever was one of the first manifestations of COVID-19 in 87.8% of cases.

Symptoms were reported by checking off specific complaints listed on the smart phone application (see Methods). In those who were test positive the comparisons of symptoms reported by those who were test positive with controls revealed 7 symptoms that demonstrated a specificity of 0.8 or higher for COVID-19: chills (0.848), stuffy nose (0.864), loss of smell and taste (0.953)), headache (0.802), trouble breathing (0.978), fatigue (0.867), and diarrhea (0.957). (Figure 2)

**Figure 2.**
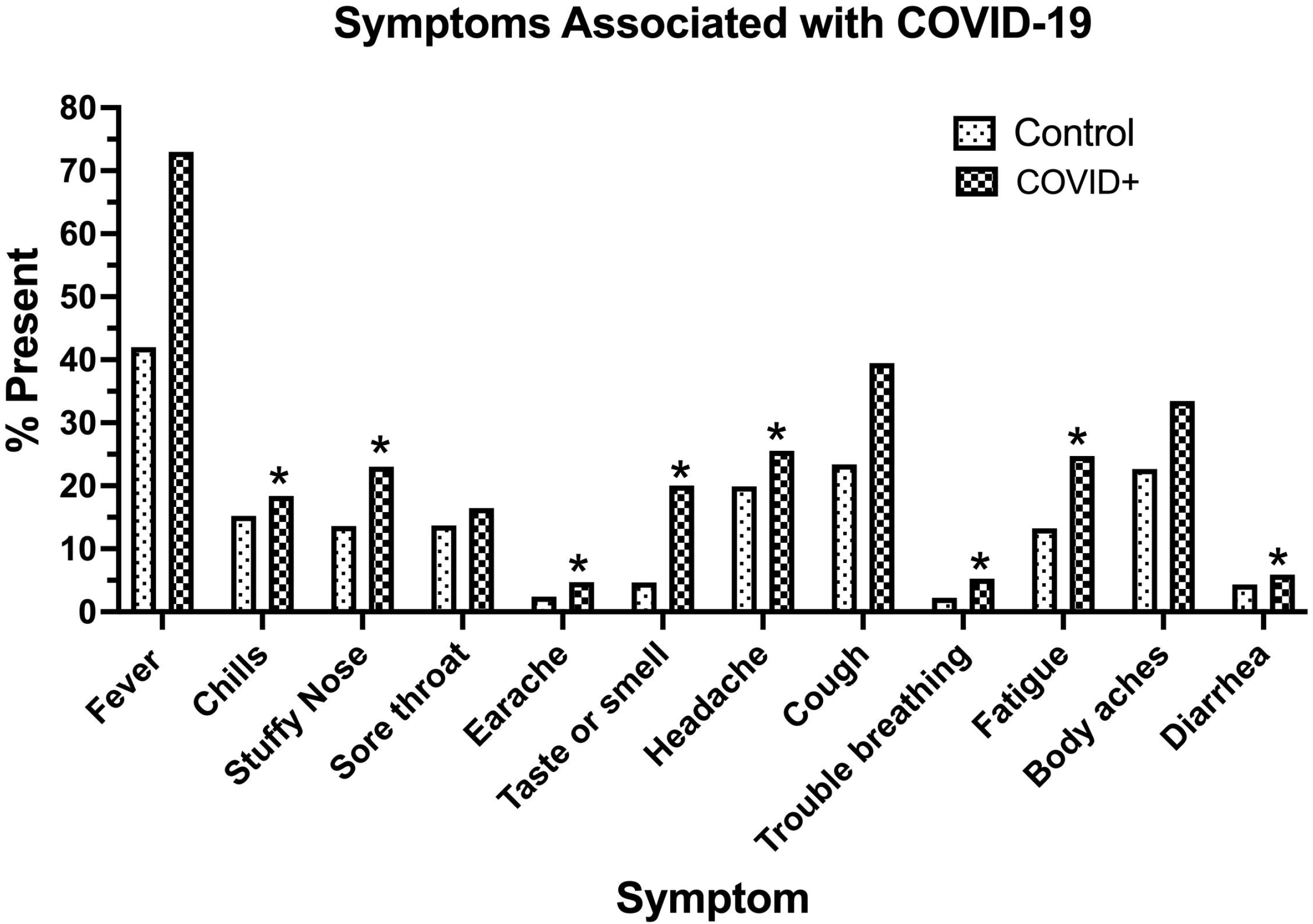
Symptoms Associated with COVID-19. The percentage of the total symptoms reported by the Control (total symptoms 1312) and COVID-19+ (total 4537) populations were calculated for each symptom. Controls had an average of 1.68 symptoms and COVID-19+ 2.44 symptoms per person. Sensitivity and specificity were determined for each symptom that achieved a statistically significant higher percentage in COVID-19 patients as compared to controls and included: loss of taste and smell: sensitivity 0.20, specificity 0.95; stuffy nose: sens. 0.23 spec. 0.86; fatigue: sens. 0.25, spec. 0.87; body aches: sens. 0.35, spec. 0.77; cough: sens. 0.39, spec. 0.77; headache: sens. 0.25 spec. 0.80; earache: sen. 0.05 spec. 0.98; chills: sens 0.18, spec 0.85; trouble breathing: sens. 0.05, spec. 0.95 and diarrhea: sens. 0.06, spec. 0.95. ** p < 0.0001 (Earache p = 0.0002) * p < 0.05

Logistic regression analysis revealed that fever combined with individual symptoms substantially increased the odds ratio for having COVID-19, loss of smell and taste being particularly high. Other predictors with high odds ratios were fever and fatigue, and fever and trouble breathing, fever and chills, fever and stuffy nose, and fever and diarrhea. (See Table 3)

**Table 3.** Logistic regression analysis of fever and specific COVID-19 symptoms.

| Symptom or sign | Odds ratio | 2.5% | 97.5% | P value | Sensitivity |
| --- | --- | --- | --- | --- | --- |
| Fever | 1.572 | 1.339 | 1.844 | < 0.0001 | 0.63 |
| Fever + Chills | 3.572 | 3.006 | 4.245 | < 0.0001 | 0.34 |
| Fever + stuffy nose | 3.624 | 2.958 | 4.441 | < 0.0001 | 0.33 |
| Fever + loss of taste and smell | 10.373 | 7.815 | 13.769 | < 0.0001 | 0.35 |
| Fever + Headache | 2.585 | 2.162 | 3.090 | < 0.0001 | 0.41 |
| Fever + Cough | 3.572 | 3.006 | 4.245 | < 0.0001 | 0.53 |
| Fever + trouble breathing | 4.219 | 2.784 | 6.394 | < 0.0001 | 0.65 |
| Fever + Fatigue | 4.098 | 3.358 | 5.000 | < 0.0001 | 0.40 |
| Fever + body aches | 2.900 | 2.440 | 3.447 | < 0.0001 | 0.49 |
| Fever + diarrhea | 3.838 | 2.772 | 5.315 | < 0.0001 | 0.66 |

In our test positive population 15.4% experienced no symptoms. In this asymptomatic group fever was less frequently detected, being reported in 46.1% (84/182) of all asymptomatic patients and in 50.9% (27/53) of asymptomatic individuals who monitored their temperature for over 72 hours.

## Discussion

### When host cells

become infected by the original SARS-CoV-2 strain, the levels of virus in saliva and nasal mucous can increase to 100 million – 1 trillion particles per ml over the first 3-10 days of infection.^10^ Because each cell is estimated to produce 100 viral particles, this suggests that 10^6^ to 10^10^ respiratory epithelial cells, dendritic cells and macrophages are serving as viral factories within a single host and each of these cells is releasing cytokines generating a strong signal to increase body core temperature.^10^ Subsequent variants, particularly the delta variant replicate more rapidly and produce virions at 10-1,000 x higher levels. Therefore, fever would be expected to be an early and frequent manifestation of all active SARS-CoV-2 infections. However, the literature to date does not fulfill this expectation. As shown in Table 4 the detection of fever in published COVID-19 cases has been extremely variable, ranging from 2% in a homeless population in Boston^29^ to 99% in hospitalized patients from China.^24^

**Table 4.**
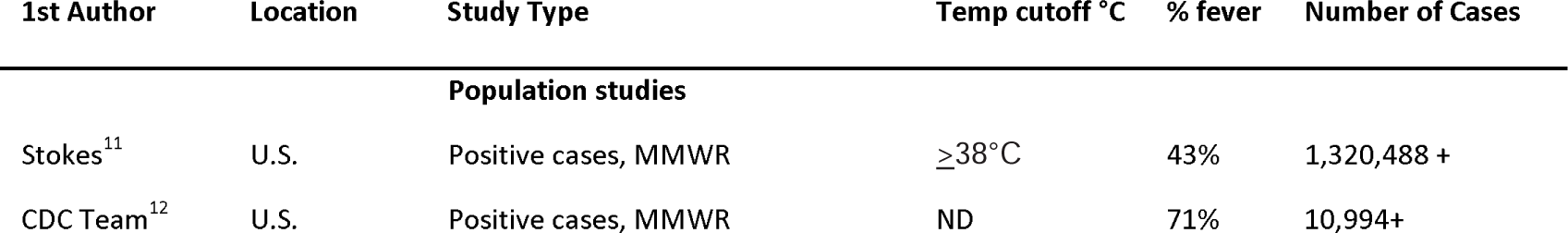
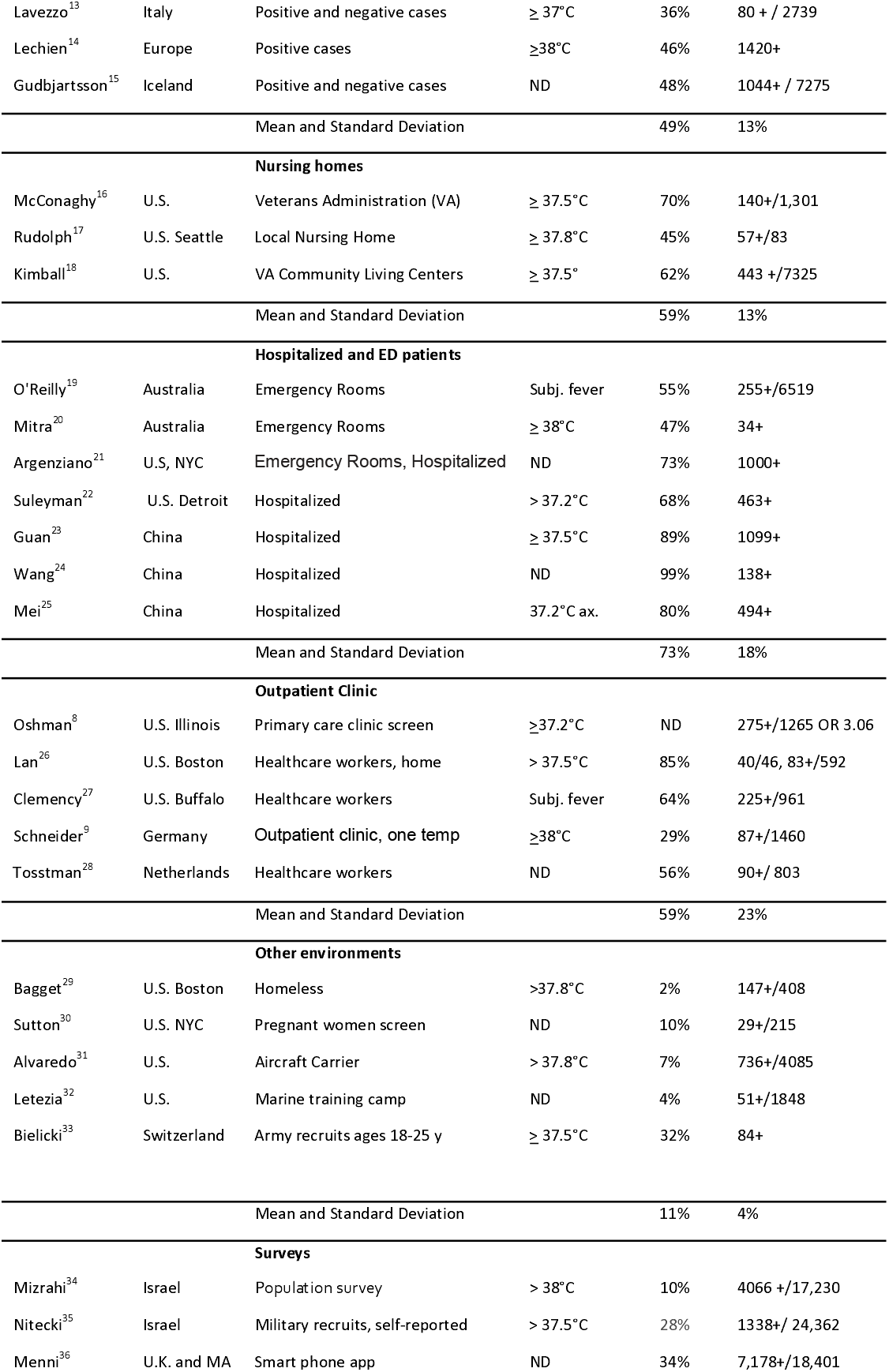
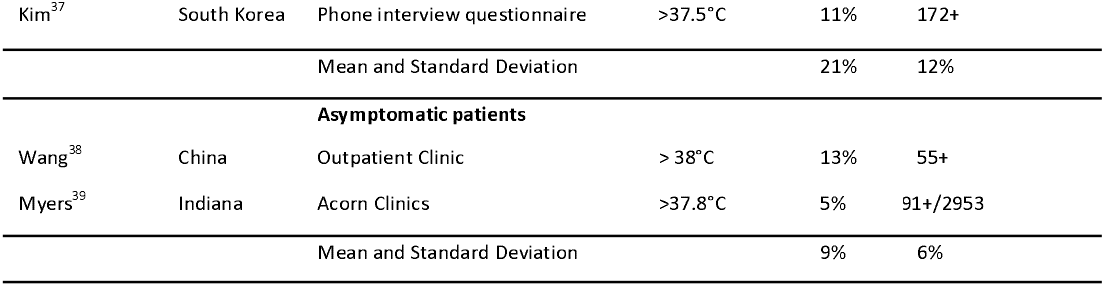
Studies of Fever in COVID-19 Positive Patients.

The lowest percentage of SARS-CoV-2 positive individuals were observed in those who had no symptoms, mean 9%, however the sample sizes were small.^38,39^ In our larger series fever was detected in 46% of asymptomatic patients. Absence of fever in these cases is likely to reflect the mild clinical presentation due to minimal organ involvement. A low prevalence was also reported in the homeless^29^, pregnant women screened on an obstetric ward^30^, and military recruits on an aircraft carrier^31^, and in military training camps^32,33^. The possible explanations for the failure to mount or detect fever could be malnutrition in the homeless, the immunosuppressive effects of pregnancy in obstetric patients, and the young age of military recruits as well as the possibility of less accurate measures of body temperature in this very mobile and active population. Large phone and paper surveys have also yielded a low prevalence of fever (mean 21%) demonstrating the poor sensitivity of this approach for documenting the symptoms and signs of COVID-19.

Large population studies have also yielded a variable prevalence of fever (36%-71%), the highest prevalence of 71% being reported in a CDC survey of over 10,000 U.S. adults who had tested positive.^12^ Another CDC report of over 1 million adults reported a fever prevalence of 43%,^11^ a percentage similar to large surveys from Iceland of 42-48%^15^and a smaller survey from Italy (36%).^13^

The highest level of fever detection has been observed in hospitalized patients and patients visiting the emergency rooms (mean 73%) closely approximating the prevalence of fever in our study. Those seeking emergency care and hospitalization are more likely to be symptomatic, to have more severe disease and to undergo multiple core temperature measurements.

Assisted living facilities and nursing homes studies have yielded moderate variations in fever prevalence (35-70%). Among residents in Veterans Administration sponsored community living centers 62% of test positive patients had fever defined as > 37.5°C, while a cutoff of 38°C identified only 24% of cases.^17^ In a study of Veterans Administration nursing homes 70% of COVID-19 patient had fever defined as a temperature of > 37.2°C.^16^ In a large outbreak of COVID-19 in a Seattle nursing home at the time of testing only 35% had symptoms including fever, however, over the ensuing week, 71% of asymptomatic patients developed fever (> 37.8°).^40^

Finally studies of outpatients have yielded a highly variable prevalence of fever in test positive individuals (29-85%). A health care worker outpatient survey found fever in 56% of those who tested positive. Among those who monitored their temperature at home 85% reported fever (defined as a temperature of > 37.5°C).^26^ Two other health care worker outpatient surveys yielded similar percentages of fever associated with positive tests (56% and 64%). One outpatient study measured a single body temperature on arrival to the clinic and detected fever (defined as temp of > 38°C) in only 29% of cases.^9^

There are several explanations for the marked variation in the prevalence of fever. First studies were conducted in different settings with different age groups. Secondly the definition of fever was variable, lower prevalence being observed for higher fever thresholds, and third, different methods were employed to identify fever included prospective direct observations, medical record reviews, retrospective surveys, and large population data bases. In addition, many of the studies focused on multiple characteristics of COVID-19 and except for two studies^20,33^ did not primarily focus on fever. Furthermore, the two studies that focused on fever consisted of very small samples raising concerns about the validity of their findings. ^20,33^

Our investigation primarily focused on the prevalence and timing of fever in SARS-CoV-2 positive individuals and analyzed over 80,000 temperature determinations. Temperature measurements were oral and were conducted at home. As observed for the outpatient survey of health care workers who measured their temperatures at home^26^, we found a high prevalence of fever, 73% when temperature was monitored for over 72 hours. We broadened our definition of fever to take into account the fact that core body temperature varies by age^7^ and defined fever as a rise of 1° C or 1.8°F in core temperature in addition to a fixed oral temperature value 37.6° C or 99.7° F. This additional criterion increased identification of fever by 28%. It should be emphasized that a single temperature measurement proved to be an insensitive screening tool only detecting fever in 30.3% of COVD-19 positive patients, a percentage comparable to that found in a large primary care clinic study that used a single temperature measurement (29%).^9^

There are several limitations to our findings. First, we analyzed a convenience sample that may not be applicable to all socioeconomic groups and these data cannot infer cause-effect due to selection bias factors; however, the survey did include a broad geographic distribution, a wide age range and a prolonged home monitoring period. Secondly COVID-19 positive tests were self-reported and may underestimate the number of positive tests. We suspect our control populations may have contained some SARS-CoV-2 infected individuals, and this condition would be expected to underestimate the specificity of fever for detecting the onset of COVID-19.

A third concern is the low specificity of fever. In comparison to a matched control population, the specificity of fever for detecting COVID-19 ranged from 0.62-0.73. However, the low specificity of fever is to be expected, and determining the etiology of fever is one of the most frequent reasons for Infectious Disease consultation.^41^ Fever serves as a nonspecific warning of possible infection and should trigger a more complete history, exam, and the ordering of specific tests to clarify the etiology. However, in the setting of a high incidence of COVID-19 the presence of fever has a higher likelihood of reflecting the onset of this disease.

As shown in Figure 3, we propose a simple management algorithm for management of individuals home monitoring for fever. First and most important for preventing spread in the workplace, school or household, the individual with fever should be immediately isolated. A symptom checklist can then be filled out and if fever is accompanied by one or more symptoms that are associated with an odds ratio of 3.5 or higher for having SARS-CoV-2 infection (loss of taste and smell, trouble breathing, fatigue, diarrhea, chills, stuffy nose or cough), particularly in resource limited environments, the individual can be presumed to have COVID-19 and continue isolation. In environments where rapid antigen tests are readily available this test can be used to confirm the diagnosis. If fever is not accompanied by these symptoms ideally an RTPCR or rapid antigen test should be performed to confirm or exclude SARS-CoV-2 infection. In individuals without fever or symptoms the probability of COVID-19 is sufficiently low except in high prevalence areas that testing will be of low yield. These individuals are unlikely to be infected and do not require isolation but should continue to monitor their temperature.

**Figure 3.**
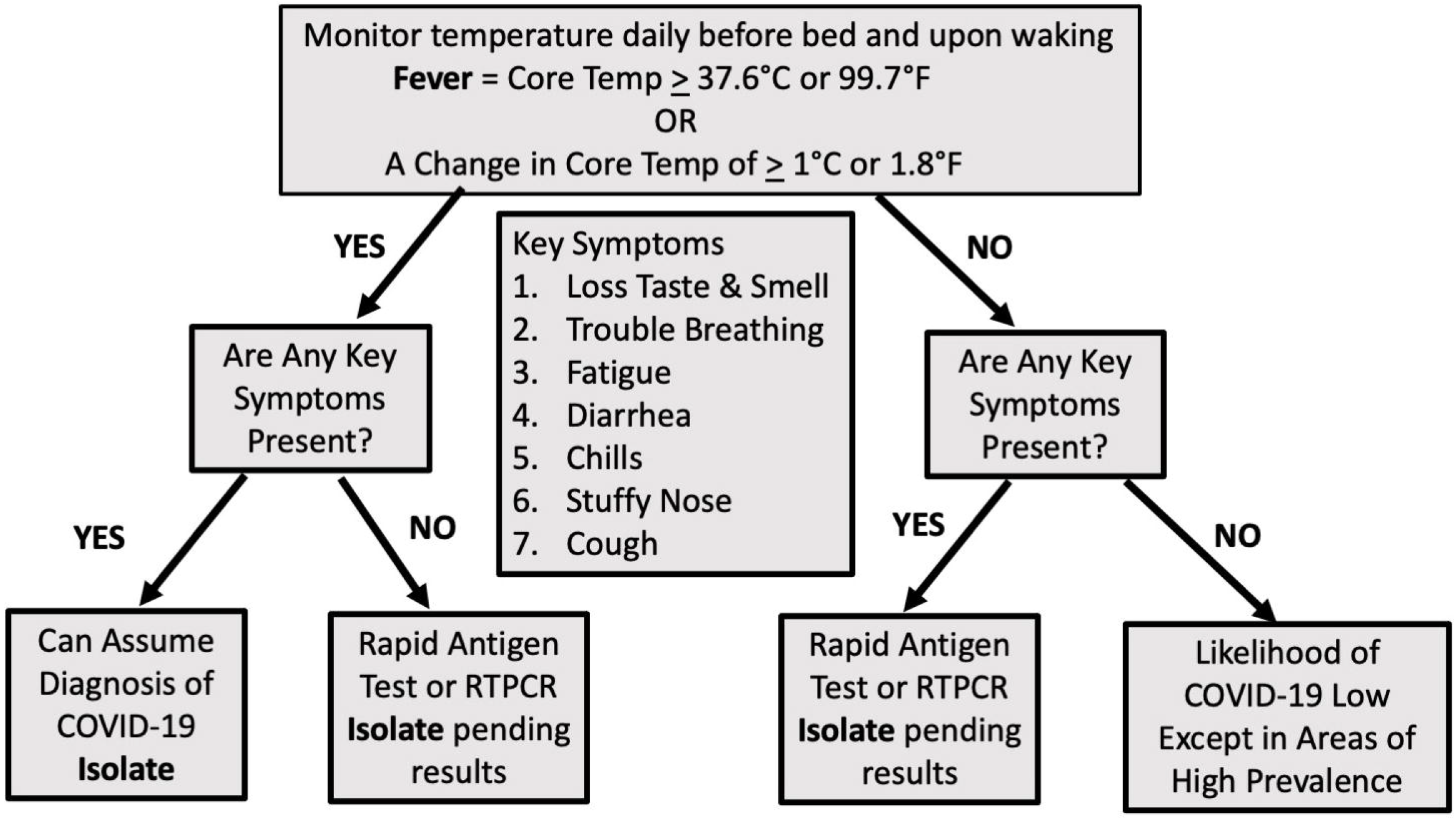
Recommended Algorithm for Monitoring Fever, Isolation and Testing in Individuals at Risk for Contracting COVID-19. (See text for details)

In conclusion, fever is an early and common sign of COVID-19 that we recommend be used as a preliminary screen before more specific RTPCR and antigen testing. This approach is inexpensive, convenient, would allow continuous monitoring for the onset of disease, and would encourage timely isolation. If widely implemented this approach has the potential to markedly reduce the spread of infection, particularly in households and other closed environments.

## Data Availability

All data is stored in secure Red Cap files at the University of Florida and has been de-identified.

## Authors’ contributions

Justin Kim compiled and organized the data using excel selection algorithms, assisted in writing the results, and reviewed the manuscript. Marcus Threadcraft assisted in analyzing the data, created the results tables, and reviewed the manuscript. Wei Xue and Sijie Yue conducted the logistic regression analysis and assisted in writing the methods and results. Richard Wenzel assisted in the research design, reviewed and edited the manuscript and assisted with data analysis. Fred Southwick – primarily designed the study, supervised the data analysis, reviewed the literature, and primarily wrote the manuscript.

## Ethics committee approval

University of Florida IRB1 approved as exempt (UF IRB# 202003028).

## Acknowledgements

We thank Inder Singh and Amy Daitch of Kinsa Inc. for providing the de-identified temperature data files.

